# Prevalence and Correlates of Ideal Cardiovascular Health among Ugandan Adolescents: A Cross-Sectional Study

**DOI:** 10.64898/2026.06.13.26355572

**Authors:** Levicatus Mugenyi, Simple Ouma, Ivan Namakoola, Arthur Namara, Isaac Samuel Kintu, Francis Xavier Namugera, Sumayiya Nalubega, Sanula Nanozi, Faith Tumuhairwe, Dorothy Mirembe, Charles Ssekyanzi, Costella M. Tindyebwa, Martin Muddu, Flavia Zalwango, Mathias Akugizibwe, Moreen Chaka Namulundu, Perez Nicholas Ochanda, Eleanor Namusoke, Mina Nakawuka, Gerald Mutungi, Rachel King, Moffat Nyirenda, David Moore, Josephine Birungi

## Abstract

**Introduction:** Cardiovascular disease (CVD) risk factors often emerge during adolescence and track into adulthood, yet data on cardiovascular health (CVH) in sub-Saharan Africa remain limited. We assessed the prevalence and correlates of ideal CVH among Ugandan adolescents.

**Methods:** We analysed baseline data of adolescents enrolled in a cluster-randomised controlled trial being conducted in urban (Kampala) and rural (Jinja) districts of Uganda. In this study, Ideal CVH was defined as meeting “ideal” status of 5-7 of the American Heart Association’s Life’s Simple 7 metrics. Random-effects logistic regression was used to identify factors associated with ideal CVH, accounting for village-level clustering.

**Results:** We recruited 1316 participants with a mean age of 13.2 years, of whom 58.1% were female. Overall, the prevalence of ideal CVH was 66.8% (95% CI: 64.2% - 69.3%). The prevalence was higher in Jinja (74.4%, 95%CI: 70.9% - 77.7%) than Kampala (59.6%, 95%CI: 55.8%-63.2%) and the difference was evident (p<0.001). Male adolescents had higher odds of ideal CVH than females in both rural (aOR=1.55, 95%CI: 1.05-2.29) and urban (aOR=1.90, 95%CI: 1.38-2.63) settings. Increasing age and higher education level were associated with lower odds of ideal CVH in both settings, likely reflecting age-related behavioural changes.

**Conclusion:** More than half of Ugandan adolescents have ideal CVH, with disparities by sex, age, and urbanisation. These findings suggest that cardiovascular health declines during adolescence and highlight the need for early, targeted interventions, particularly among female and urban adolescents.

## Introduction

Cardiovascular diseases (CVDs) represent a major global health challenge, accounting for approximately 18 million deaths annually, with over three-quarters occurring in low- and middle-income countries (LMICs) (1,2). While CVDs typically manifest in adulthood, there is increasing evidence that their origins lie earlier in life, with risk factors emerging during childhood and adolescence and tracking into later life (3–5).

Adolescence represents a critical period for the establishment of health behaviours that influence long-term cardiovascular risk, including diet, physical activity, and tobacco use. The American Heart Association’s concept of ideal cardiovascular health (CVH), defined by seven modifiable metrics (Life’s Simple 7), provides a useful framework for assessing overall cardiovascular risk profiles (6). Evidence from high-income countries shows that maintaining ideal CVH in early life is associated with substantially lower risk of CVD and improved cardiometabolic outcomes in adulthood (7,8). However, data on CVH among adolescents in LMICs, particularly in sub-Saharan Africa, remain scarce.

Countries in sub-Saharan Africa, including Uganda, are undergoing rapid epidemiological and demographic transitions, characterised by urbanisation, changing dietary patterns, and reduced physical activity and growing burden of non-communicable diseases (NCDs) alongside persistent infectious diseases (9). Recent surveys indicate rising prevalence of CVD risk factors such as hypertension, obesity, and physical inactivity among Ugandan adults (10,11). Most available data focus on adults, with limited understanding of how cardiovascular risk is distributed and develops during adolescence.

Importantly, cardiovascular health may differ substantially between urban and rural populations due to variations in lifestyle, socio-economic conditions, and environmental exposures (12–14). Understanding these differences is essential for designing context-specific prevention strategies, particularly in settings experiencing rapid urbanisation.

In this study, we assessed the prevalence of ideal cardiovascular health among adolescents in Uganda using the Life’s Simple 7 framework and examined socio-demographic and behavioural factors associated with ideal CVH in both urban and rural settings. By identifying early patterns of cardiovascular health, this study aims to inform targeted interventions to prevent the development of CVD in this population.

## Methods

### Study design and setting

This study uses baseline data from a cluster-randomised controlled trial evaluating the effectiveness of a family centred approach in the enhancement of lifestyle change and behavioural modification for prevention of cardiovascular diseases among adolescents and their families in Uganda (“FaCe-D” study). The trial is being conducted in Kampala and Jinja districts representing urban and rural settings in Uganda, respectively. Kampala, is a capital city of Uganda located in the central region of the country, and Jinja is located about 80 kilometers east of Kampala. The study sites included one urban division (Nakawa) in Kampala and two rural sub-counties (Budondo and Mafubira) in Jinja.

### Study population

From 5^th^ July to 19^th^ October 2025, we enrolled participants from 32 selected villages called clusters (16 in Kampala district and 16 in Jinja district). We worked with Village Health Teams (VHTs) and local council one (LC1) leaders to generate a list of villages within the selected sites that had approximately 40-50 households with each having at least one adolescent. Villages which did not share borders (to avoid contamination of the trial intervention) were considered for selection. The inclusion criteria for the participants were: 1) willingness of the household head or parent to provide permission or consent, 2) adolescents aged 10-19 years, and 3) readiness to stay in the study village for the next 12 months. Adolescents were excluded if they were in boarding school or planning to join boarding school, pregnant or lactating, confirmed to be receiving treatment for a chronic condition such as CVD, diabetes, asthma, cancer or mental illness, not able to provide assent or consent, and those whose caregivers/parents did not provide consent (for adolescent aged <18 years). For each household, one adolescent who met the inclusion and none of the exclusion criteria was selected (or randomly selected if there were more than one adolescent in the household) and invited to participate in the study during baseline data collection. The targeted sample size was 1,280 participants obtained assuming a prevalence of ideal CVH among adolescents of 53% based on the South African study (15), 90% power of detecting 10% unit difference in the prevalence between the intervention groups, 5% alpha, 32 clusters of size 40, and an intra-class correlation coefficient of 0.005. At the baseline, a total of 1,316 participants was recruited and included in the analysis of this paper.

### Participant consent and assent

Prior to recruitment into the study, trained research team staff obtained written informed consent from parents or caregivers (or household heads) and adolescent aged 18-19 years, as well as assent from adolescents aged 10-17 years. Assent was obtained only from adolescents aged 10-17 years whose parents or caregivers provided consent. The study received research ethics approval from the Uganda Virus Research Institute, the Uganda National Council for Science and Technology, and the London School of Hygiene and Tropical Medicine. The trial is registered at clinicaltrials.gov, NCT07265453 (https://clinicaltrials.gov/study/NCT07265453).

### stakeholder involvement

During the formative phase, prior to baseline data collection, the stakeholders were involved in the adaptation of the proposed family-based intervention (16) to the Ugandan context. This included stakeholders’ meetings, informant interviews and focus group discussions. The stakeholders included household heads, primary household food preparers, community health workers, VHT members, policymakers at Ugandan Ministry of Health (MoH) and at district levels, non-government organisation (NGO) representatives, religious and cultural leaders, and health care providers at primary health facilities located in the study sites. VHTs provide an important link between households and the formal healthcare system, while adolescents identified as peer champions support in promoting behavioural change among adolescents.

### Data collection

Trial participants were asked to complete an interviewer-administered questionnaire on tablets using RedCap software. Data included socio-demographic, socio-economic and household-level characteristics. In addition, data were collected on the life’s simple 7 metrics of cardiovascular health including smoking, body mass index (BMI), physical activity, healthy diet, blood pressure, total cholesterol, and fasting blood glucose. The specific questions on the 7 metrices, the possible responses, and how the responses were used to derive cardiovascular ideal situation for each metric are given in the supplementary Table 1. Also, data were collected on depression, anxiety, and sleep quality as measures of mental health to assess their associations with cardiovascular health. Only baseline data were used for this paper.

### Data management

Trained research assistants checked the data for completeness daily before syncing it to the RedCap database located on the MRC/UVRI and LSHTM Uganda Research Unit institutional computer server. The trial statistician exported the data to Stata version 19 software for further cleaning and analysis.

### Outcome measure

The outcome for this paper was ideal cardiovascular health (ideal CVH) defined as proportion of adolescents who scored 5-7 of the life’s simple 7 metrics. For each metric, the adolescents were scored 1 if they met the cut-off for ideal cardiovascular health and 0 if otherwise. A total score (for cardiovascular health) was obtained by summing up the 7 individual metric scores. Table 1 summarizes the cut-off points for ideal cardiovascular health that were considered for each metric.

**Table 1.**
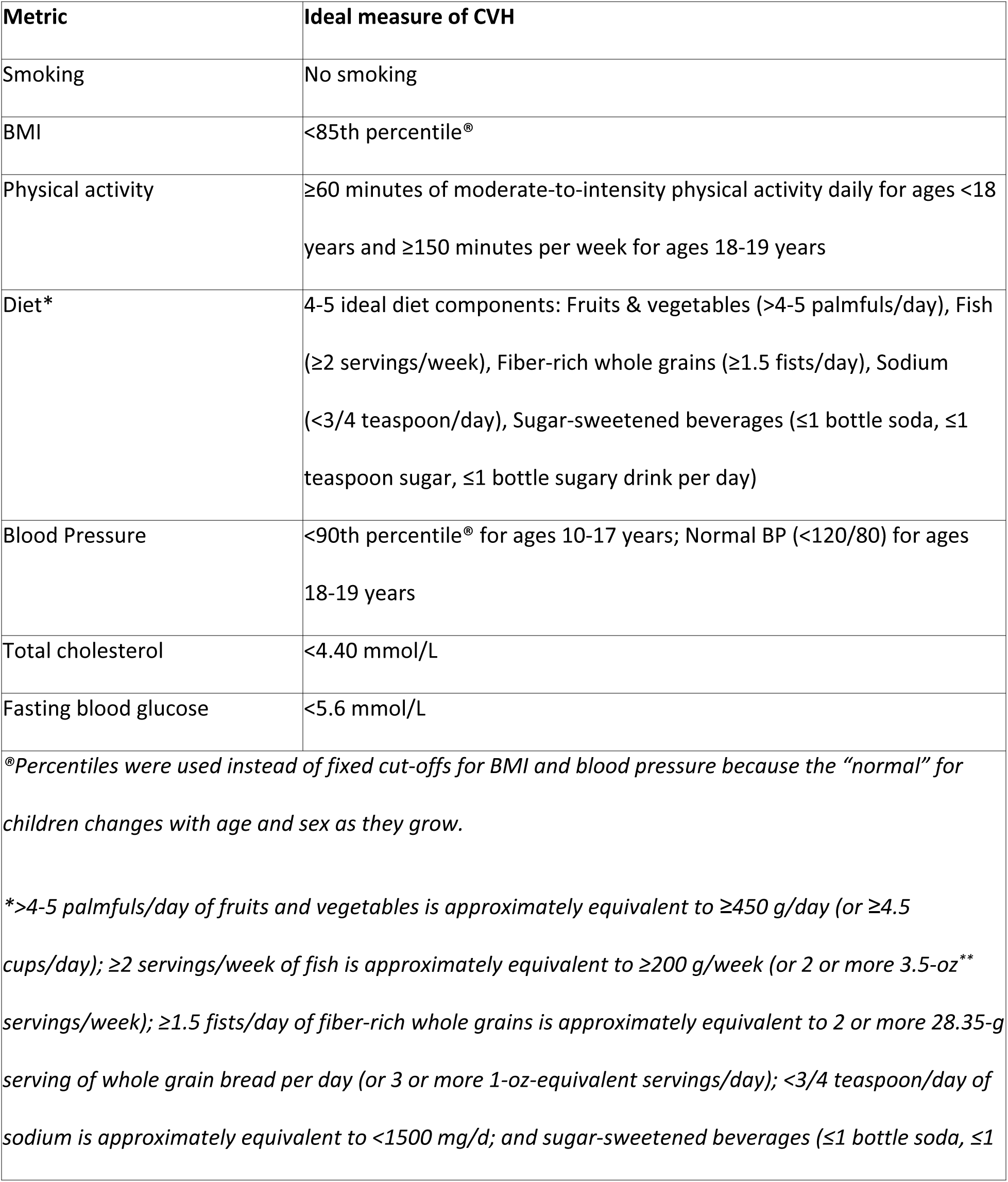
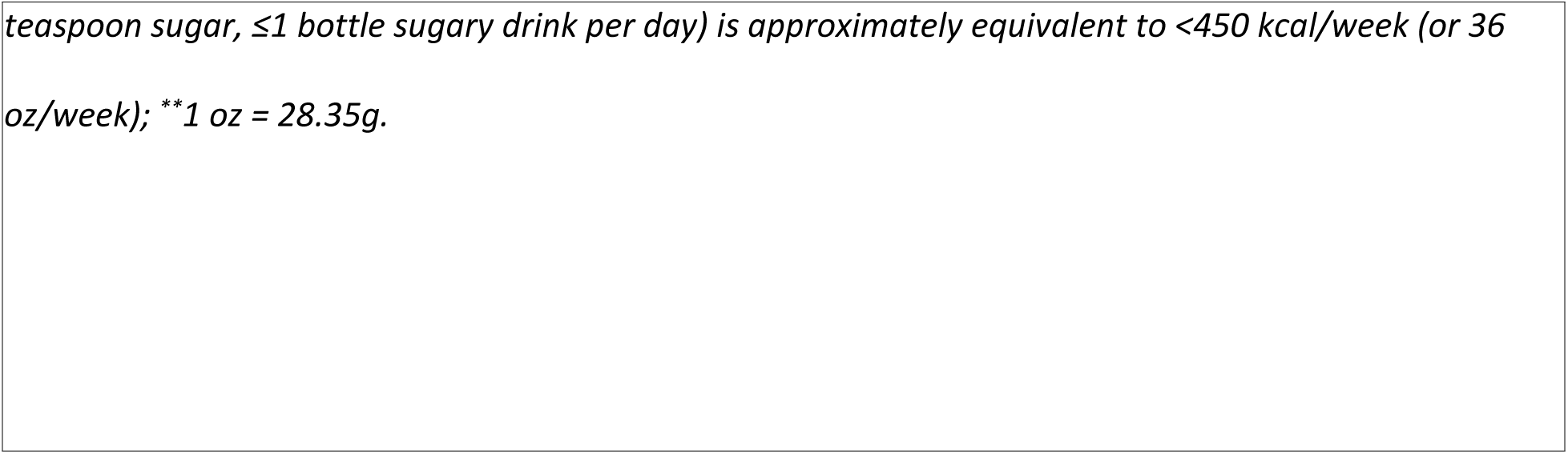
The life’s simple 7 framework customized to our study.

| Metric | Ideal measure of CVH |
| --- | --- |
| Smoking | No smoking |
| BMI | <85th percentile<sup>®</sup> |
| Physical activity | ≥60 minutes of moderate-to-intensity physical activity daily for ages <18 years and ≥150 minutes per week for ages 18-19 years |
| Diet* | 4-5 ideal diet components: Fruits & vegetables (>4-5 palmfuls/day), Fish (≥2 servings/week), Fiber-rich whole grains (≥1.5 fists/day), Sodium (<3/4 teaspoon/day), Sugar-sweetened beverages (≤1 bottle soda, ≤1 teaspoon sugar, ≤1 bottle sugary drink per day) |
| Blood Pressure | <90th percentile<sup>®</sup> for ages 10-17 years; Normal BP (<120/80) for ages 18-19 years |
| Total cholesterol | <4.40 mmol/L |
| Fasting blood glucose | <5.6 mmol/L |
| <sup>®</sup>Percentiles were used instead of fixed cut-offs for BMI and blood pressure because the “normal” for children changes with age and sex as they grow. |  |
| *>4-5 palmfuls/day of fruits and vegetables is approximately equivalent to ≥450 g/day (or ≥4.5 cups/day); ≥2 servings/week of fish is approximately equivalent to ≥200 g/week (or 2 or more 3.5-oz** servings/week); ≥1.5 fists/day of fiber-rich whole grains is approximately equivalent to 2 or more 28.35-g serving of whole grain bread per day (or 3 or more 1-oz-equivalent servings/day); <3/4 teaspoon/day of sodium is approximately equivalent to <1500 mg/d; and sugar-sweetened beverages (≤1 bottle soda, ≤1 |  |

*teaspoon sugar, $\leq 1$ bottle sugary drink per day) is approximately equivalent to $<450$ kcal/week (or 36 oz/week); \*\*1 oz = 28.35g.*

### Statistical analyses

We used Stata version 19 software and R version 4.4.1 to perform all the analyses. Participants’ characteristics were summarized by study site using frequencies and proportions for categorical data and means (standard deviation) for numerical data. In addition to the outcome variable (ideal CVH), the other variables used in the analysis included urbanisation, age, sex, ethnicity, family size, schooling status (in and out of school), highest education level, household monthly income, sleep quality, depression scores, alcohol use, cooking style, and reporting any illness in the past month.

To determine factors associated with ideal CVH, we applied a hierarchical conceptual framework with non-modifiable factors at Level 1 and modifiable factors at Level 2 (see Fig 1). The framework demonstrates that Level 1 factors (e.g., age, sex, and ethnicity) can influence Level 2 factors (e.g., urbanisation, socio-economic, psychological, behavioural and lifestyle, and clinical factors), which may in turn affect the ideal CVH in adolescents. In this framework, we further show that ideal CVH is derived from the life’s simple 7 metrics. Due to clustering of adolescents in the villages, we used a random-effects (RE) logistic regression to determine factors associated with ideal CVH. To quantify the associations, odds ratios (ORs) and 95% confidence intervals (CIs) were estimated. Each variable was assessed for its association with the outcome in a simple RE logistic regression (bivariable analysis) and the strength of evidence determined using the Wald test statistic. Results from the bivariable analysis are presented as unadjusted estimates.

**Fig 1.**
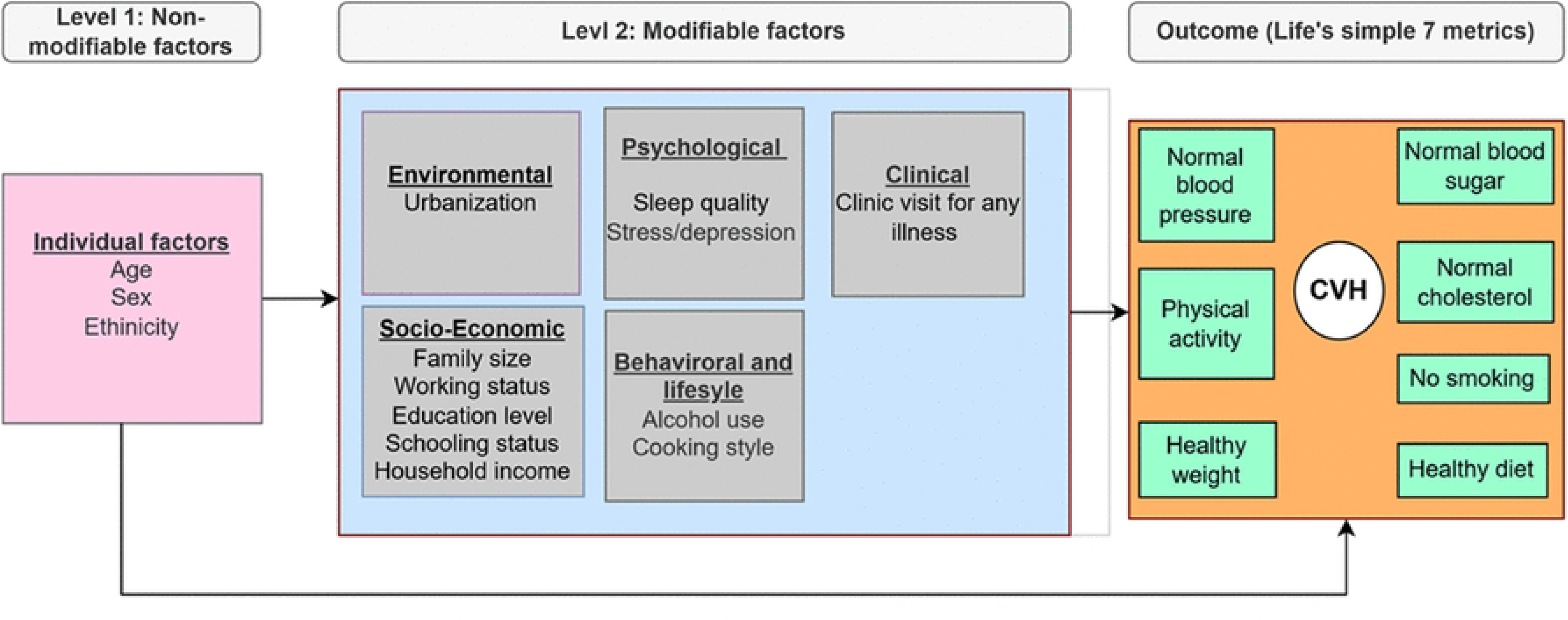
Conceptual framework showing plausible relationships between baseline factors and the cardiovascular health.

For multivariable analysis, first, we included level 1 factors into the RE logistic regression model and variables potentially associated with ideal CVH (p<0.1) were retained and adjusted for each other. Then, level 2 factors were included in the model adjusted for each other, and for the factors retained at Level 1. Variables that were associated with the outcome were retained in the final model and results presented as adjusted estimates. Global Wald test was used to determine factors associated with ideal CVH. The intra-cluster correlation (ICC) in cardiovascular health among adolescents within the same village was estimated using the final model.

## Results

### Socio-demographic characteristics

A total of 1,316 adolescents were recruited and provided data for analysis. All participants had data available on all baseline variables. The mean age was 13.2 (standard deviation, SD=2.6) years, 58.1% were females, and about half (51.3%) were enrolled from Kampala district (urban setting). The largest proportion (47.9%) were of Basoga/Bagwere ethnic group and more than half (57.3%) were from households of size ranging from 4-6 people. A larger proportion (29.7%) of the adolescents live in households with an average monthly income of less than 30 USD. Details of the participants’ characteristics stratified by district (urban vs rural) are given in Table 2.

**Table 2:** Baseline Socio-demographic factors by district.

|  | Kampala (urban) | Jinja (rural) | Total |
| --- | --- | --- | --- |
|  | (N=675) | (N=641) | (N=1,316) |
| **Mean age in years (SD)** | 13.2 (2.7) | 13.1 (2.5) | 13.2 (2.6) |
| **Age group in years** |  |  |  |
| 10-11 | 232 (34.4%) | 204 (31.8%) | 436 (33.1%) |
| 12-13 | 166 (24.6%) | 188 (29.3%) | 354 (26.9%) |
| 14-15 | 138 (20.4%) | 119 (18.6%) | 257 (19.5%) |
| 16-17 | 82 (12.1%) | 87 (13.6%) | 169 (12.8%) |
| 18+ | 57 (8.4%) | 43 (6.7%) | 100 (7.6%) |
| **Sex** |  |  |  |
| Female | 380 (56.3%) | 385 (60.1%) | 765 (58.1%) |
| Male | 295 (43.7%) | 256 (39.9%) | 551 (41.9%) |
| **Level of education** |  |  |  |
| Primary | 526 (77.9%) | 506 (78.9%) | 1,032 (78.4%) |
| Secondary | 138 (20.4%) | 123 (19.2%) | 261 (19.8%) |
| Tertiary | 11 (1.6%) | 12 (1.9%) | 23 (1.7%) |
| **Currently working** |  |  |  |
| Yes | 46 (6.8%) | 4 (0.6%) | 50 (3.8%) |
| No | 629 (93.2%) | 637 (99.4%) | 1,266 (96.2%) |
| **Ethnic group** |  |  |  |
| Acholi/Langi | 40 (5.9%) | 0 (0.0%) | 40 (3.0%) |
| Alur | 22 (3.3%) | 7 (1.1%) | 29 (2.2%) |
| Baganda | 183 (27.1%) | 49 (7.6%) | 232 (17.6%) |
| Banyankole/Bakiga | 77 (11.4%) | 8 (1.2%) | 85 (6.5%) |
| Basoga/Bagwere | 89 (13.2%) | 541 (84.4%) | 630 (47.9%) |
| Banyoro/Batoro | 35 (5.2%) | 0 (0.0%) | 35 (2.7%) |
| Itesot/Japhadola | 85 (12.6%) | 16 (2.5%) | 101 (7.7%) |
| Lugbara | 33 (4.9%) | 3 (0.5%) | 36 (2.7%) |
| Samia | 15 (2.2%) | 8 (1.2%) | 23 (1.7%) |
| Bagisu/Banyole | 67 (9.9%) | 6 (0.9%) | 73 (5.5%) |
| Other East Africa | 23 (3.4%) | 2 (0.3%) | 25 (1.9%) |
| Other | 6 (0.9%) | 1 (0.2%) | 7 (0.5%) |
| **Household size** |  |  |  |
| 1-4 | 263 (39.0%) | 115 (17.9%) | 378 (28.7%) |
| 4-6 | 358 (53.1%) | 396 (61.8%) | 754 (57.3%) |
| 7+ | 53 (7.9%) | 130 (20.3%) | 183 (13.9%) |
| **Monthly income (USD)\*** |  |  |  |
| <30 | 96 (14.2%) | 295 (46.0%) | 391 (29.7%) |
| 30-55 | 142 (21.0%) | 143 (22.3%) | 285 (21.7%) |
| 56-96 | 186 (27.6%) | 128 (20.0%) | 314 (23.9%) |
| 97+ | 251 (37.2%) | 75 (11.7%) | 326 (24.8%) |

*\*Exchange rate: 1USD=UGX 3700. Quartiles were used to categorize monthly income*

### Cardiovascular Health Score

First, we explored the adolescents’ scores on the life’s simple 7 metrics. The score (0–7) corresponds to the number of metrics on which the adolescent met the cardiovascular health cut-off, with 0 corresponding to worst and 7 to best cardiovascular health. Fig 2 shows the percentage distribution of the adolescents’ scores grouped by study site. Most (45.0%) of the adolescents scored 5 (out of 7) points with the percentage slightly higher in Jinja than Kampala (46.8% vs 43.3%). The lowest ideal cardiovascular health scores were on the diet (3.9%) and physical activity (33.1%) metrics. The percentages meeting the ideal thresholds were consistently higher among rural than urban adolescents for all metrices, except for blood pressure. Fig 3 shows the percentage of adolescents who met the cardiovascular health cut-off for each metric grouped by study site.

**Fig 2.**
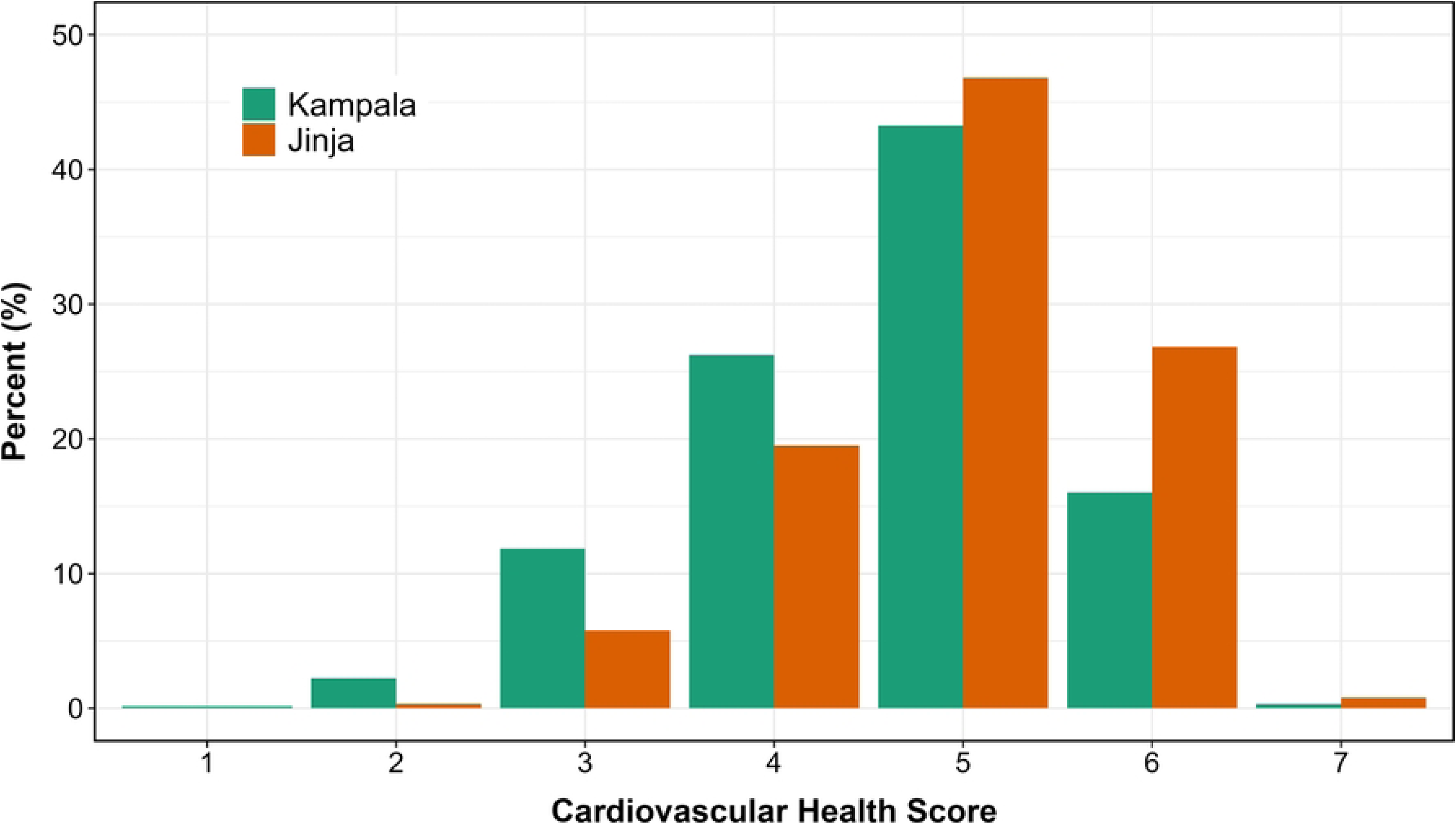
Percentage distribution of cardiovascular health score (–) based on life’s simple 7 metrics.

**Fig 3.**
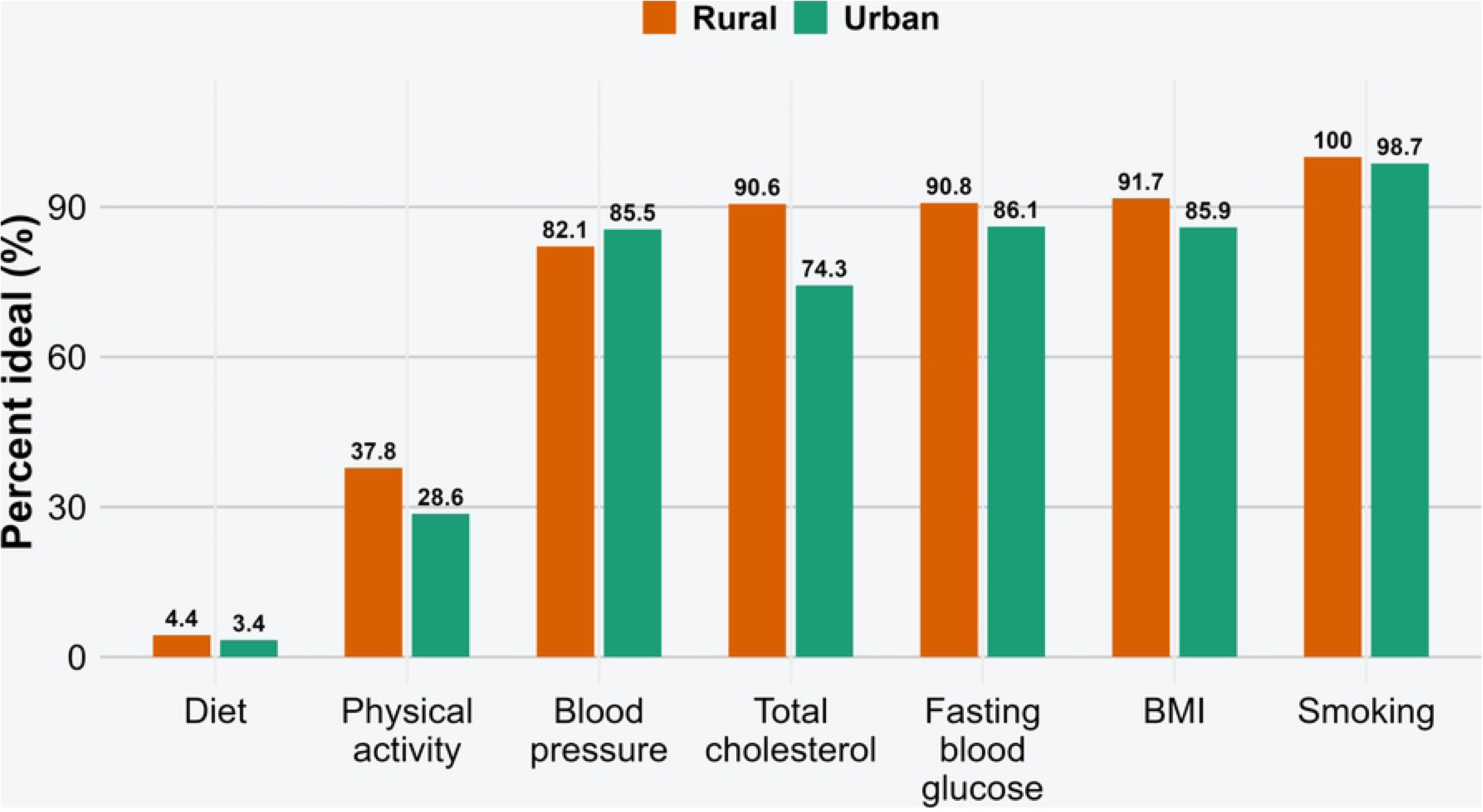
Percentage distribution of participants that were ideal on life’s simple 7 metrics.

### Prevalence of Ideal Cardiovascular Health

Overall, the prevalence of ideal CVH (score 5-7) was 66.8% (95% CI: 64.2% - 69.3%). The prevalence was higher in Jinja (74.4%, 95%CI: 70.9% - 77.7%) than Kampala (59.6%, 95%CI: 55.8%-63.2%) and the difference was evident (p<0.001).

### Factors Associated with Ideal Cardiovascular Health

Due to differences in cardiovascular health scores between districts (urbanisation effect) as presented in the above paragraph (and in Fig 2 and Fig 3), we present results on factors associated with ideal CVH stratified by district (urban and rural).

Table 3 presents results on factors associated with ideal CVH among adolescents in a rural setting of Jinja district. Results show that after accounting for village-level clustering and other factors, ideal CVH in a rural setting was associated with age, sex, ethnic group, and education level. The prevalence of ideal CVH decreased with increasing age (aOR=0.82, 95%CI: 0.77-0.88; p<0.001), and it was higher among male than female adolescents (79.7% vs 70.9%; adjusted odds ratio (aOR=1.55, 95%CI: 1.05-2.29; p=0.027. In Jinja, the prevalence of ideal CVH was higher among adolescents in other ethnic groups than the indigenous group (Basoga/Bagwere) (83.4% vs 73.2%), though the difference was not significant (p=0.094). Consistent with age effect, the prevalence with ideal CVH was lower among adolescents with tertiary than primary education (25.0% vs 79.4%; aOR=0.15, 95%CI: 0.04-0.62, p=0.009). Also, there was a borderline an association between visiting a clinic due to any illness in the past month and cardiovascular health (aOR=0.32, 95%CI: 0.09-1.12, p=0.074). The factors that were not associated with ideal CVH in a rural setting included sleep quality, alcohol use, household size, working status, schooling status, household income, cooking style (e.g., use of cooking oils), and depression/stress. Results on factors not associated with ideal CVH are presented in supplementary Table 2.

**Table 3.** Baseline factors associated with ideal cardiovascular health among adolescents in a rural setting (Jinja district)

|  | Ideal CVH |  | Unadjusted |  | Adjusted |  |
| --- | --- | --- | --- | --- | --- | --- |
|  | Total | n (%) | OR (95% CI) | P | OR (95% CI) | P |
| **Level 1 Factors (Non-modifiable)** |  |  |  |  |  |  |
| **Age in years** |  |  | 0.82 (0.76, 0.88) | <0.001 | 0.82 (0.77, 0.88) | <0.001 |
| **Sex** |  |  |  |  |  |  |
| Female | 385 | 273 (70.9) | 1.0 |  | 1.0 |  |
| Male | 256 | 204 (79.7) | 1.63 (1.11, 2.38) | 0.012 | 1.55 (1.05, 2.29) | 0.027 |
| **Level 2 Factors (Modifiable)** |  |  |  |  |  |  |
| **Level of education** |  |  |  |  |  |  |
| Primary | 506 | 402 (79.4) | 1.0 |  | 1.0 |  |
| Secondary | 123 | 72 (58.5) | 0.36 (0.23, 0.55) | <0.001 | 0.62 (0.34, 1.12) | 0.114 |
| Tertiary | 12 | 3 (25.0) | 0.08 (0.02, 0.32) | <0.001 | 0.15 (0.04, 0.62) | 0.009 |
| **Visited** |  |  |  |  |  |  |
| **clinic/hospital for** |  |  |  |  |  |  |
| **any illness in the** |  |  |  |  |  |  |
| **past month** |  |  |  |  |  |  |
| Yes | 31 | 28 (90.3) | 1.0 |  | 1.0 |  |
| No | 610 | 449 (73.6) | 0.30 (0.09, 1.02) | 0.055 | 0.32 (0.09, 1.12) | 0.074 |

Table 4 presents results for factors associated with ideal CVH among adolescents in an urban setting (Kampala). Similar to the rural setting, there was strong evidence for association of ideal CVH with sex, age, and education among adolescents in the urban setting. Male adolescents had a higher proportion with ideal CVH compared to their female counterparts (68.5% vs 52.6%, aOR=1.90, 95%CI: 1.38-2.63; p<0.001), and the prevalence of ideal CVH decreased with increasing age (aOR=0.91, 95%CI: 0.86-0.97, p=0.002). Furthermore, the results show evidence for reduced ideal CVH among adolescents with increased level of education (trend test: p=0.007). For example, the prevalence of ideal CVH was lower among adolescents in secondary than primary level (45.7% vs 63.7%, aOR=0.49, 95%CI: 0.29-0.84, p=0.009), and lower among those with tertiary education than primary (36.4% vs 63.7%, aOR=0.33, 95%CI: 0.09-1.28, p=0.110). Also, though a weak evidence, the prevalence of ideal CVH was lower among out-of-school adolescents than those that were currently in school (48.1% vs 61.1%, aOR=0.60, 95%CI: 0.35-1.03, p=0.066). The factors that were not associated with ideal CVH in an urban setting included ethnic group, sleep quality, alcohol use, household size, working status, household income, cooking style (e.g., use of cooking oils), depression/stress, and reported illness. Results for factors that were not associated with ideal CVH are presented in supplementary Table 3.

**Table 4.**
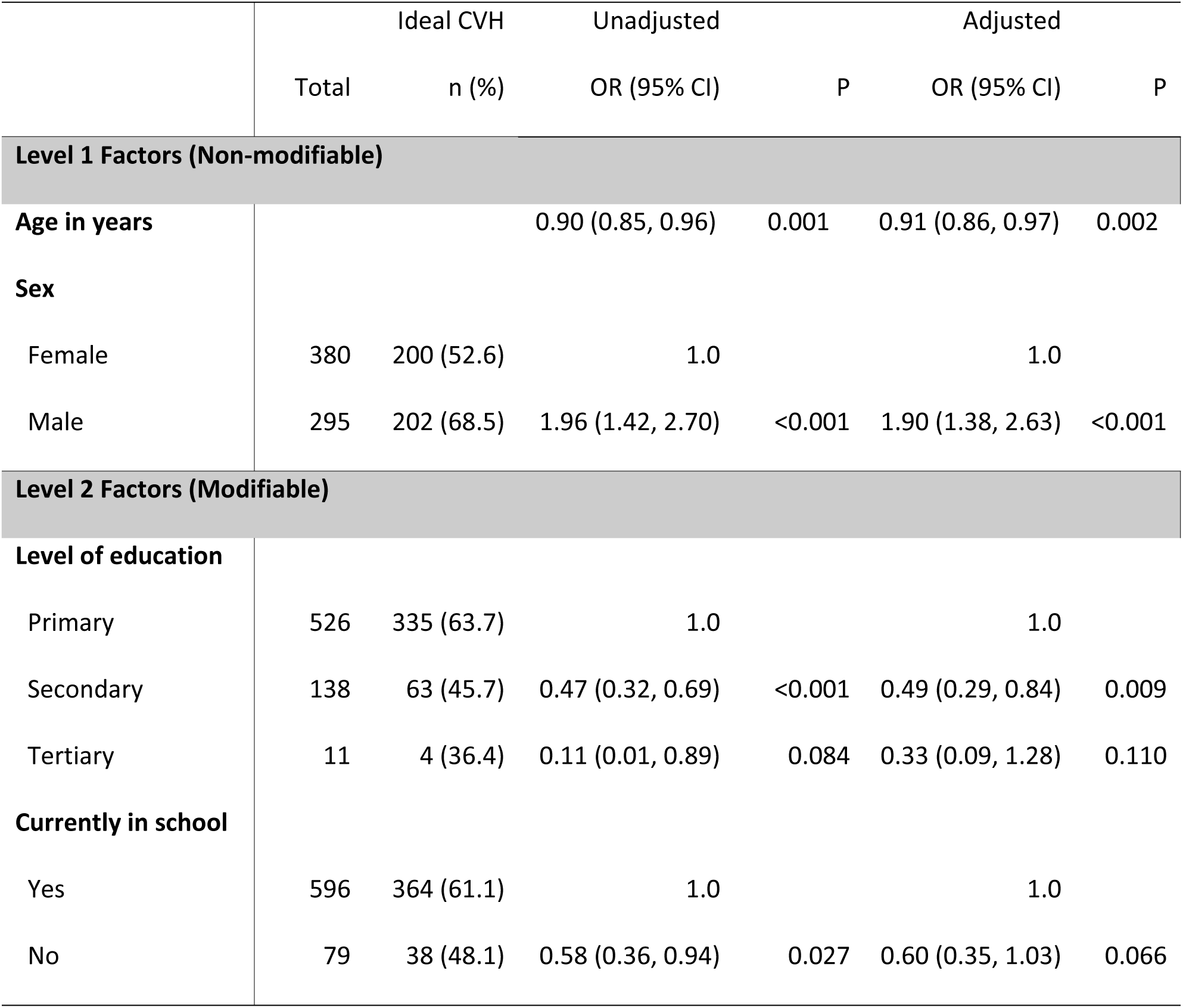
Baseline factors associated with ideal cardiovascular health among adolescents in an urban setting (Kampala district)

## Discussion

In this study of over 1,300 adolescents in Uganda, we found that two-thirds met criteria for ideal cardiovascular health (CVH), with marked disparities by sex, age, and urbanisation. The key drivers for reduced CVH were poor diet and physical inactivity. These findings highlight that suboptimal cardiovascular health is already common in early life in this setting, underscoring adolescence as a critical window for prevention of future cardiovascular disease.

### Prevalence of Ideal Cardiovascular Health

The overall prevalence of ideal CVH (66.8%) suggests that a substantial proportion of adolescents are already accumulating cardiovascular risk. While this estimate falls within ranges reported in high-income settings(17,18), the implications in the Ugandan context may be more profound given the rapid epidemiological transition and limited health system capacity to manage chronic disease. Importantly, our findings suggest that deterioration in CVH begins early, reinforcing a life-course perspective in which exposures during adolescence shape long-term cardiometabolic risk.

A key finding was the substantially higher prevalence of ideal CVH in rural compared to urban adolescents. This pattern is consistent with the hypothesis that urbanisation is associated with adverse shifts in health behaviours, including reduced physical activity, increased consumption of processed foods, and more sedentary lifestyles (19). In contrast, rural adolescents may benefit from more physically active daily routines and less exposure to highly processed diets. However, these apparent advantages may diminish over time as rural environments undergo similar transitions. These findings highlight the need to address emerging cardiovascular risk in rapidly urbanising settings, while also anticipating future risk in rural populations.

### Sex Differences in Cardiovascular Health

We observed a consistent sex disparity, with male adolescents more likely to have ideal CVH than females in both settings. This difference likely reflects a combination of behavioural, social, and biological factors. In many contexts, boys may have greater opportunities for physical activity through sports and outdoor activities, whereas girls may face sociocultural constraints or competing domestic responsibilities (20). Additionally, gendered patterns of diet, body image, and physical activity may emerge during adolescence and influence cardiovascular risk profiles. These findings point to the importance of gender-sensitive interventions that address structural and social barriers to healthy behaviours among female adolescents.

### Age-Related Decline in Cardiovascular Health

The inverse association between age and ideal CVH suggests that cardiovascular health declines during adolescence. This may reflect increasing autonomy over lifestyle choices, including diet and physical activity, as well as greater exposure to unhealthy environments, particularly in urban settings (21). The observed association between higher educational level and lower odds of ideal CVH likely reflects, at least in part, confounding by age, as older adolescents are more likely to have progressed further in education. It may also capture behavioural transitions associated with schooling environments, such as reduced physical activity or changes in dietary patterns. These findings should therefore be interpreted cautiously and not as evidence that education itself is detrimental to cardiovascular health.

### Strengths and Limitations

Our study has several strengths, including a large sample size, inclusion of both urban and rural populations, and use of a standardised framework for assessing CVH. However, several limitations should be considered. First, the cross-sectional design precludes causal inference and limits our ability to assess trajectories of cardiovascular health over time. Second, some components of the CVH metrics, particularly diet and physical activity, were based on self-report and may be subject to measurement error and social desirability bias. In addition, the adaptation of Life’s Simple 7 metrics to the local context, while necessary, may limit comparability with other studies. Third, residual confounding cannot be excluded, particularly for socio-behavioural factors that were not fully captured.

### Implications for Policy and Practice

Despite these limitations, our findings have important implications. They suggest that interventions to promote cardiovascular health should begin early in adolescence and be tailored to specific high-risk groups, particularly female and urban adolescents. School- and community-based strategies that promote physical activity, healthy diets, and broader health literacy may be especially relevant. Furthermore, longitudinal follow-up of this cohort will be important to understand how cardiovascular health evolves over time and to identify critical windows for intervention.

### Conclusions

This study demonstrated that two-thirds of Ugandan adolescents have ideal cardiovascular health, with significant disparities by sex, age, and urbanisation. Male adolescents, younger adolescents, and those in rural settings had higher prevalence of ideal cardiovascular health. The declining cardiovascular health with age highlights the urgency of adolescent-focused prevention strategies. These findings provide important baseline data for the FaCe-D trial and offer insights for designing culturally appropriate cardiovascular health promotion programmes for adolescents in Uganda and similar low-resource settings in sub-Saharan Africa. Future research should focus on identifying effective intervention strategies and understanding the mechanisms underlying observed disparities to inform evidence-based prevention efforts.

## Data Availability

The dataset used and analyzed during the current study is available from the corresponding author on reasonable request

## List of abbreviations

AHA: American Heart Association
BMI: Body Mass Index
CI: Confidence Interval
CVD: Cardiovascular Disease
CVH: Cardiovascular Health
FaCe-D: Family Centred Approach for Cardiovascular Disease Prevention
ICC: Intra-Cluster Correlation
ICVH: Ideal Cardiovascular Health
IQR: Interquartile Range
LC1: Local Council One
LMICs: Low- and Middle-Income Countries
LSHTM: London School of Hygiene and Tropical Medicine
MoH: Ministry of Health
MRC: Medical Research Council
NCDs: Non-Communicable Diseases
NGO: Non-Governmental Organisation
OR: Odds Ratio
RE: Random Effects
SD: Standard Deviation
UNCST: Uganda National Council for Science and Technology
UVRI: Uganda Virus Research Institute
VHT: Village Health Team

## Ethics approval and consent to participate

Ethical approval was obtained from the Uganda Virus Research Institute (UVRI) Research Ethics Committee (reference GC/127/991) and the Uganda National Council for Science and Technology (UNCST) (reference HS3682ES), and the London School of Hygiene and Tropical Medicine (LSHTM) Research Ethics Committee (reference 30553). Informed consent to participate was obtained from all participants.

## Availability of data and materials

The dataset used and analysed during the current study is available from the corresponding author on reasonable request.

## Competing interests

The authors declare no financial and non-financial competing interests.

## Transparency statement

The manuscript is an honest, accurate, and transparent account of the study being reported; and no important aspects of the study have been omitted.

## Funding statement

This work was funded by the Global Alliance for Chronic Disease through the Canadian Institutes for Health Research (Grant Ref. GR027342). The funder had no role in the design of the study; in the collection, analyses, or interpretation of the data; in the writing of the article; or in the decision to publish the results.

## Authors’ contributions

LM conceived the study, conducted the analysis, and wrote the first draft of the manuscript. All authors contributed to the study design, interpretation of findings, and critical revision of the manuscript. All authors read and approved the final manuscript.

## Acknowledgements

We extend our sincere thanks to study participants and their families, Village Health Teams, local council leaders, the FaCe-D research team, and all stakeholders who contributed to this study. We acknowledge the support of the MRC/UVRI and LSHTM Uganda Research Unit.

## Notes

### Competing Interest Statement

The authors have declared no competing interest.

### Clinical Trial

NCT07265453

### Funding Statement

Yes

